# Identifying groups of people with similar sociobehavioural characteristics in Malawi to inform HIV interventions: a Latent Class Analysis

**DOI:** 10.1101/2019.12.26.19015560

**Authors:** Aziza Merzouki, Amanda Styles, Janne Estill, Zofia Baranczuk, Karen Petrie, Olivia Keiser

## Abstract

**Background:** Within many sub-Saharan African countries including Malawi, HIV prevalence varies widely between regions. This variability may be related to the distribution of population groups with specific sociobehavioural characteristics that influence the transmission of HIV and the uptake of prevention. In this study, we intended to identify groups of people in Malawi with similar risk profiles.

**Methods:** We used data from the Demographic and Health Survey in Malawi from 2016, and stratified the analysis by sex. We considered demographic, socio-behavioural and HIV-related variables. Using Latent Class Analysis (LCA), we identified clusters of people sharing common sociobehavioural characteristics. The optimal number of clusters was selected based on the Bayesian information criterion. We compared the proportions of individuals belonging to the different clusters across the three regions and 28 districts of Malawi.

**Results:** We found nine clusters of women and six clusters of men. Most women in the clusters with highest risk of being HIV infected were living in female-headed households and were formerly married or in a union. Among men, older men had the highest risk of being HIV infected, followed by young (20-25) single men. Generally, low HIV testing uptake correlated with lower risk of having HIV. However, rural adolescent girls had a low probability of being tested (48.7%) despite a relatively high HIV prevalence. Urban districts and Southern region had a higher percentage of high-prevalence and less tested clusters of individuals than other areas.

**Conclusions:** LCA is an efficient method to find clusters of people sharing common HIV risk profiles, identify particularly vulnerable population groups, and plan targeted interventions focusing on these groups. Tailored support, prevention and HIV testing programmes should focus particularly on female household heads, adolescent girls living in rural areas, older married men, and young men who have never been married.

**Funding:** The project was funded by the Swiss National Science Foundation (grant no 163878).

## Introduction

Diagnosis of HIV infected people is a critical step to improve access to treatment and control the HIV epidemic. In Eastern and Southern Africa, where HIV prevalence is the highest in the world [1], progress has been made to scale up HIV testing. The proportion of people living with HIV who know their status increased from 77% in 2015 to 85% in 2018[1]. In order to keep up with the progress and identify the remaining HIV infected people, testing programs should better target those who are most at risk of having HIV.

Within many sub-Saharan African (SSA) countries, HIV prevalence varies widely between regions. This heterogeneity may result from the spatial distribution of population groups with specific sociobehavioural characteristics that promote or impede the transmission of HIV and the uptake of preventive measures. Improving our understanding of the characteristics of different groups in the population, their risk of being HIV infected and their uptake of HIV testing is essential to design more targeted and tailored prevention and care programs. Clustering analysis, which is an unsupervised machine learning technique, allows us to identify similarity patterns and find hidden sub-groups of people with potentially different risk levels and drivers of having or acquiring HIV[2]. In a previous study, we investigated differences between SSA countries and identified sociobehavioural profiles that coincided with a high HIV prevalence and incidence at the country level[3]. However, this past study did not consider sub-national differences. Moreover, previous studies on high-risk groups in SSA countries mainly focused on women and excluded men[4–6]. Most of these studies also did not analyse the spatial distribution of the identified groups across the country. A recent study mapped high-risk men and women across Malawi to study whether their distribution could explain the spatial variability of HIV prevalence in the country, but the definition of high-risk behaviour was based only on the total number of lifetime sexual partners of individuals[7].

In the present study, we aimed to identify and characterize vulnerable groups who are most at risk of being HIV positive in Malawi. We used Latent Class Analysis (LCA) to identify clusters of men and women with different sociobehavioural characteristics, based on the latest Demographic and Health Survey (DHS)[8] of Malawi. We also compared the most common clusters across the three regions and 28 districts of Malawi.

## Methods

### Data

We used data from the latest available DHS (2015-2016) for Malawi. The DHS programme collects nationally representative data and covers a wide range of indicators for population and health.

We considered datasets (**Table S1**) that contained the survey responses of men (aged 15-54 years) and women (15-49), and their HIV status (positive, negative, unknown). Two researchers (AM and OK) preselected the following variables because they covered topics that could be related to the HIV epidemic: place of residence (rural; urban), age (≤19; 20 to 25; 26 to 35; >35 years), literacy (can read a complete sentence: yes; no), access to media at least once a week (yes; no), currently employed (yes; no), marital status (formerly married or lived in a union; currently married or living in a union; never married nor lived in a union), sex of household head (female; male), age at first sexual intercourse (<16; 16 to 18; 19 to 21; >21), condom use (wife justified to ask husband to use a condom when he has a sexually transmitted infection (STI); not justified), wife beating (at least one reason justifies wife beating; no reason justifies it), comprehensive correct knowledge about AIDS (yes; no), and prior HIV testing (ever been tested; never tested). We also considered the district and region of residence of respondents for mapping.

### Analysis

We conducted separate analyses for women and men. Since our study focused on sexual transmission of HIV, we excluded persons who never had sexual intercourse. We used multiple imputation by chained equations (MICE)[9,10] to complete missing data (Supplementary Material). Some implausible or inconsistent values were set to missing before imputation.

We identified clusters of people sharing common sociobehavioural characteristics using LCA[11], a multivariate technique that groups individuals with similar characteristics in latent (i.e. not directly observable) classes (clusters). We used maximum likelihood estimation to calculate the probability of an individual belonging to a cluster. We selected the number of clusters that minimized the Bayesian information criterion (BIC)[12]. We performed the final analysis based on a subset of variables identified as the most important for clustering (Supplementary Material). We selected these variables for men and women separately based on the overall variance of their categories’ probabilities across the clusters, similarly to the method by Dean and Raftery[13]. Reducing the number of variables allows us to reduce the number of clusters, thus facilitating interpretability.

We analyzed the HIV prevalence and the uptake of HIV testing in each LCA cluster. We defined HIV prevalence as the proportion of positive tests among the positive and negative tests results. We also analyzed the distribution of the clusters across regions and districts, and assessed whether it could help to explain the wide spatial variability of HIV prevalence.

We conducted the analysis using the open source R language, version 3.5.1. We used the implementation of LCA provided in the poLCA package, version 1.4.1[14,15]. The code is available on GitLab (https://gitlab.com/AzizaM/dhs_malawi_lca).

### Ethics approval

The DHS data are freely available on request. Thus, no ethical approval was needed.

## Results

The survey we used included 24,562 women and 7,478 men. After imputing incomplete data and removing respondents who never had sexual intercourse, we performed the analysis on 21,564 women and 6,379 men. The HIV status was known for 6,750 women (5,889 HIV negative; 861 HIV positive) and 5,642 men (5,168 HIV negative; 474 HIV positive).

### Women’s clusters

We found an optimal number of nine clusters for women based on seven variables: age, literacy, marital status, location of residence, sex of household head, having ever been tested for HIV, and employment (**Table 1**). Two clusters had a high HIV prevalence (*Clusters 2* and *8*). *Cluster 8* had an HIV prevalence of 37.2%, and was predominantly urban (100.0%) and literate (87.8%). *Cluster 2* had a prevalence of 21.2%, and was predominantly rural (96.6%). In comparison, other clusters had a median HIV prevalence of 10.0% (IQR 7.9%-12.3%). *Clusters 2* and *8* both had high proportions of female-headed households (100.0% and 83.2%, respectively) and most women had lived in a union before (76.2% and 59.1%, respectively).

**Table 1.**
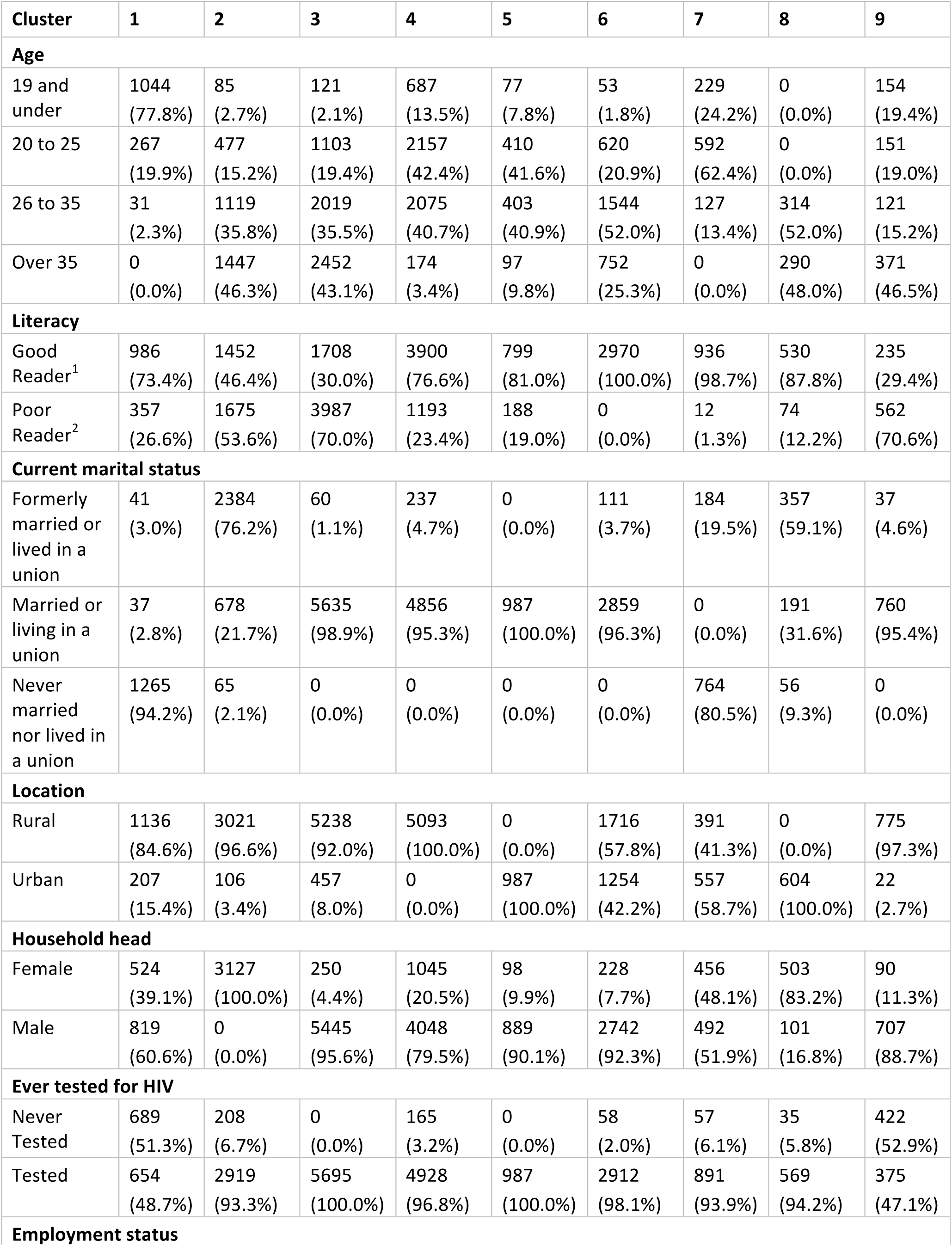
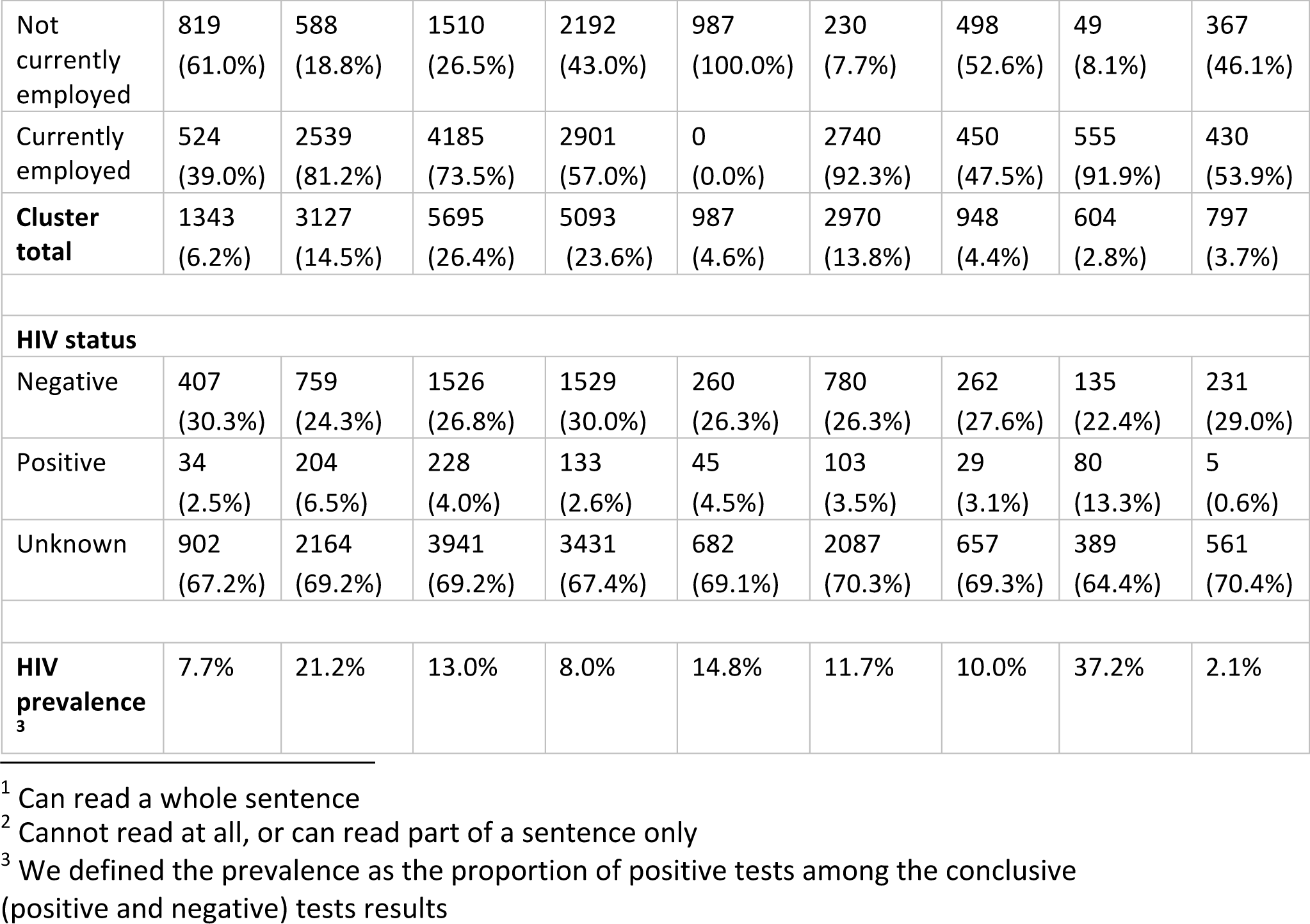
Clusters of women based on the seven most important variables: age, literacy, marital status, location of residence, sex of household head, having ever been tested for HIV and employment. Data from 21,564 women were analysed.

HIV testing uptake was relatively low in *Clusters 1* and *9* (48.7% in *Cluster 1*; 47.1% in *Cluster 9*) compared to other clusters where the median HIV testing uptake was 96.8% (IQR 94.1%-99.1%). *Cluster 9* had a low HIV prevalence (2.1%) and consisted mainly of older (46.5% over 35 years) women who lived in rural areas (97.3%) and had a low level of literacy (29.4%). These women generally lived in a union (95.4%) and their household was male-headed (88.7%). In *Cluster 1*, the HIV prevalence was higher (7.7%). This cluster included literate (73.3%), adolescent (77.8% under 20 years) girls who lived in rural (84.6%) areas and had never been in a union (94.2%). HIV testing uptake was approximately two-fold higher (96.8%) in *Cluster 4* than *Cluster 1* (48.7%), although their HIV prevalence were similar (8.0% in *Cluster 4* vs. 7.7% in *Cluster 1*). *Cluster 4* was characterized by young (83.1% aged between 20 and 35), rural (100.0%) and married (95.3%) women.

**Figure 1** and **Table S2** present the distributions of the clusters of women in the Northern, Central and Southern regions of Malawi. The Southern region included a higher percentage (7.2%) of rural, literate, adolescent girls, who had never been in a union and who had a low uptake of HIV testing (*Cluster 1*); and a lower percentage (10.8%) of well tested married women aged over 20 (*Cluster 6*) than the Central (*Cluster 1*, 6.0%; *Cluster 6*, 16.7%) and Northern (*Cluster 1*, 3.3%; *Cluster 6*, 15.1%) regions. The Southern region had also a higher percentage (17.0%) of rural divorced/widowed women with a female household head and a high HIV prevalence (*Cluster 2*) than the other regions (13.1% in the Central and 9.6% in the Northern regions). In contrast, the Northern region had a higher percentage (32.5%) of young rural married women (*Cluster 4*) with a high HIV testing uptake than the Central (21.6%) and Southern (23.2%) regions.

**Figure 1.**
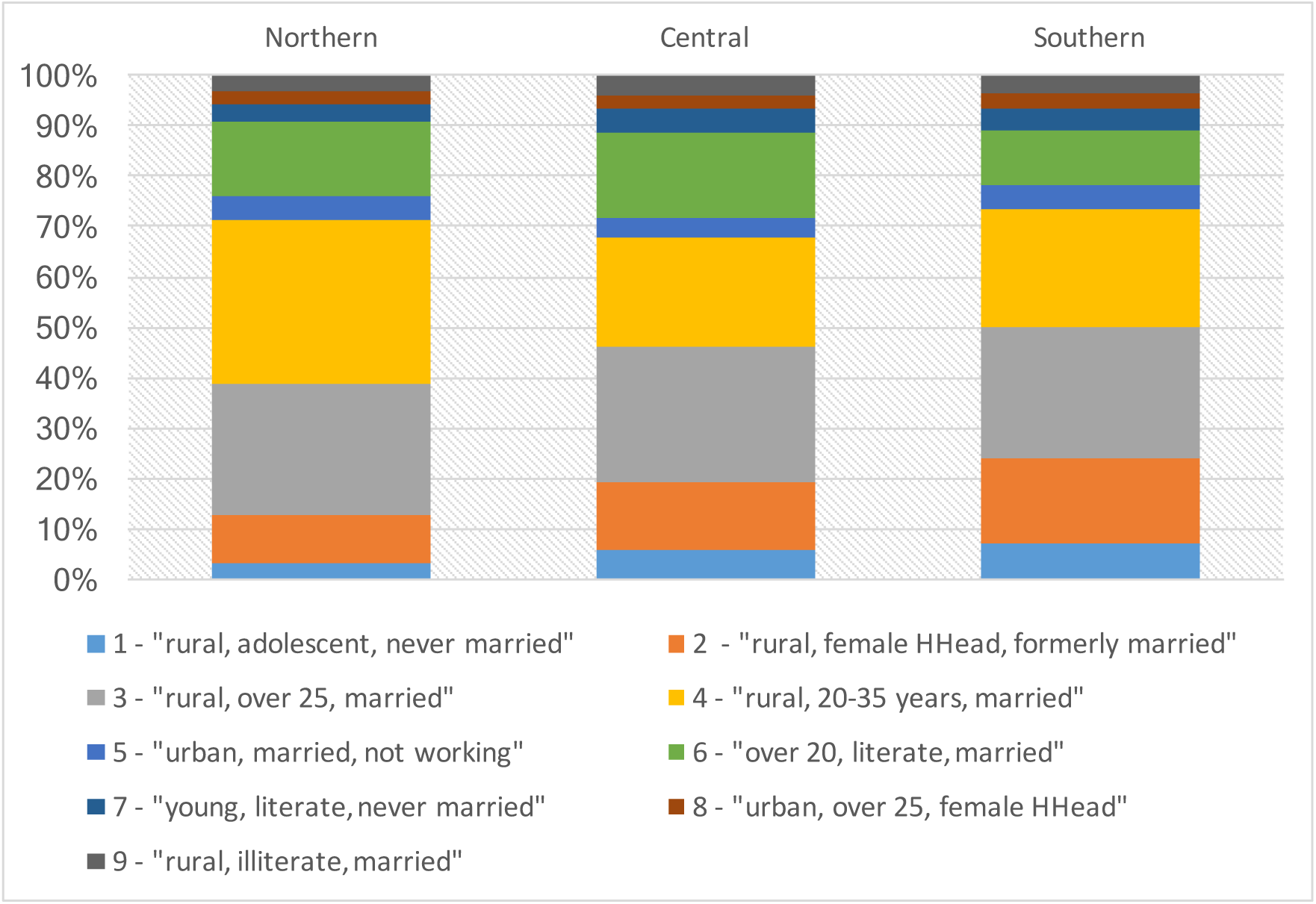
Distribution of the nine identified clusters of women in the three regions of Malawi: Northern, Central, and Southern.

The distribution of clusters of women varied also across the 28 districts of Malawi (**Table S3). Figure 2** presents the proportion of the nine clusters in five selected districts that cover all three regions of the country: Chitipa (Northern), Salima (Central), Lilongwe (Central), Blantyre (Southern) and Chikwawa (Southern region). We selected two neighbouring districts from the same region (Salima and Lilongwe in the Central region; Chikwawa and Blantyre in the Southern region) because they had remarkably different HIV prevalence (5.2% in Salima vs 9.2% in Lilongwe; 9.1% in Chikwawa vs 24.1% in Blantyre). **Table S4** and **Figure S1** present the HIV prevalence per district for women and men. In both Central and Southern regions, the districts with higher HIV prevalence (Lilongwe and Blantyre) contained the two largest cities of Malawi, and therefore were characterized by a lower level of rural clusters (*Clusters 1, 2, 3* and *4)*, and a higher percentage of more urban clusters (*Clusters 5, 6, 7* and *8*). In the Southern region, *Cluster 9*, which is 97.3% rural and has the lowest HIV prevalence, was substantially more common in Chikwawa than in Blantyre (7.2% vs 1.0%), unlike in the Central region where its proportion was similar in Salima and Lilongwe (2.4%; 3.1%). Similarly to districts with lower HIV prevalence in the Central and Southern regions of Malawi, in Chitipa (Northern region) a relatively high percentage of people belonged to rural clusters, such as *Cluster 3* (33.9%), *Cluster 4* (29.4%) and *Cluster 9* (5.6%), whereas urban clusters, such as *Cluster 5* (1.7%), *Cluster 7* (2.2%) and *Cluster 8* (1.7%), were much less common. But Chitipa had also a lower percentage of *Cluster 1* (1.7%; less tested and mainly rural with relatively high HIV prevalence, vs. 10.0% in Salima and 7.0% in Chikwawa) and a lower percentage of *Cluster 2* (7.8%; rural with high HIV prevalence and female-headed households, vs. 16.9% in Salima and 12.2% in Chikwawa). Chitipa also had a higher percentage of women from *Cluster 6* (16.1%; well tested, literate, employed women, who were living in union, with a male household head, vs. 9.6% in Salima and 5.9% in Chikwawa).

**Figure 2.**
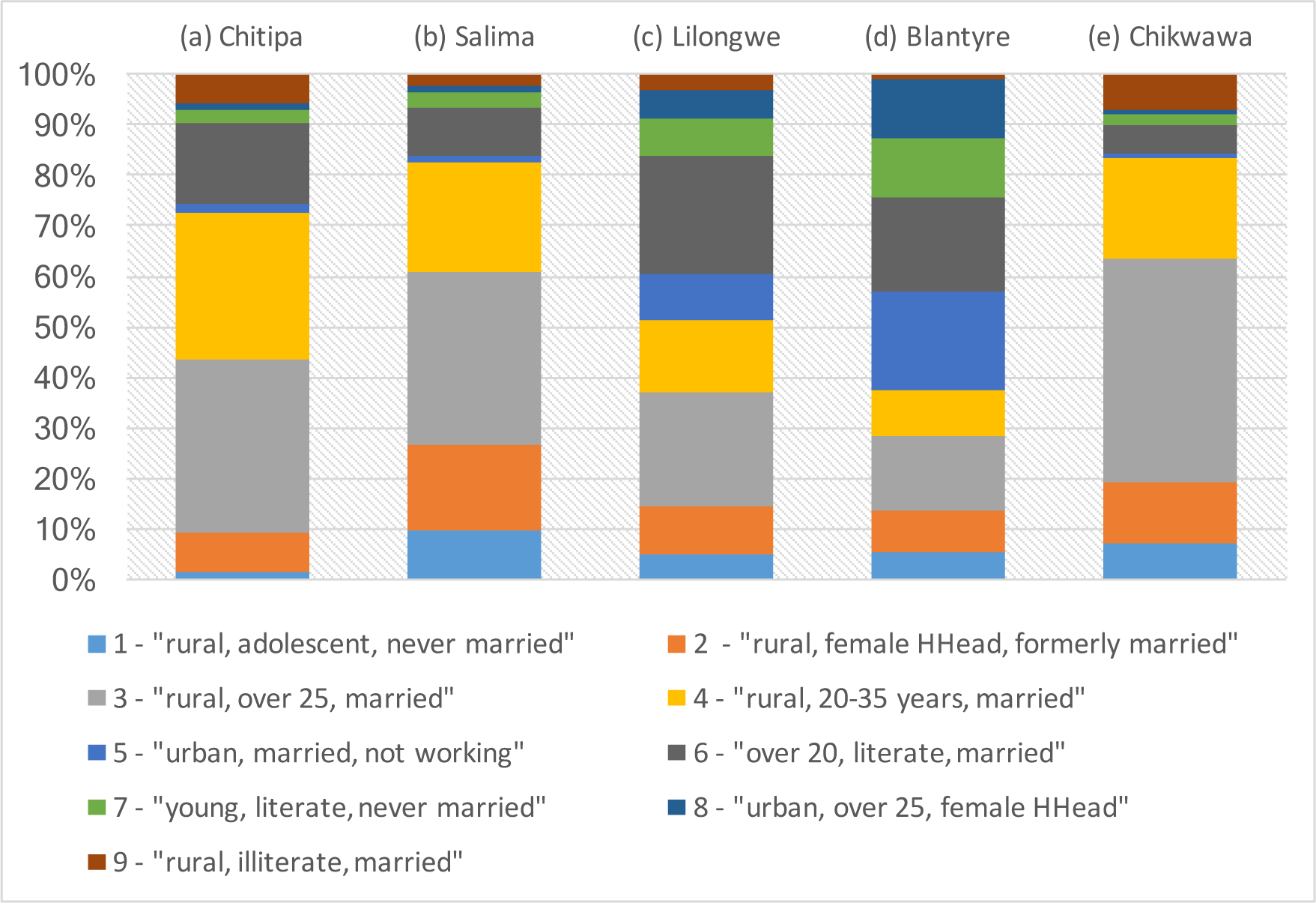
Proportion of the nine female clusters in five selected districts of Malawi: Chitipa, (b) Salima, (c) Lilongwe, (d) Blantyre, (e) Chikwawa.

### Men’s clusters

The optimal number of clusters for men was six based on 11 variables: age, age at first sex, marital status, location of residence, having ever been tested for HIV, media access, employment, literacy, sex of household head, wife beating justification, and willingness to use condom (**Table 2**). Compared to women, men had a lower HIV prevalence (8.4% vs 12.7%); and a lower uptake of HIV testing. The percentage of men who had never been tested for HIV was three times higher than for women (22.5% vs 7.6%). Two clusters of men, *Clusters 3* and *5*, had a particularly low testing uptake (52.5% and 35.0%; vs. a range of 80.5% to 92.2% in other clusters). Although literacy was substantially different between these clusters (97.7% in *Cluster 3*; 29.7% in *Cluster 5*), men in both clusters had many similar characteristics: they were mainly adolescents (77.7% under 20 years in *Cluster 3*; 70.3% in *Cluster 5*), had never lived in a union (96.0% in *Cluster 3*; 98.2% in *Cluster 5*), and lived in rural settings (80.3% in *Cluster 3*; 97.2% in *Cluster 5*). These clusters also had the lowest HIV prevalence (0.8% and 0.5%).

**Table 2.**
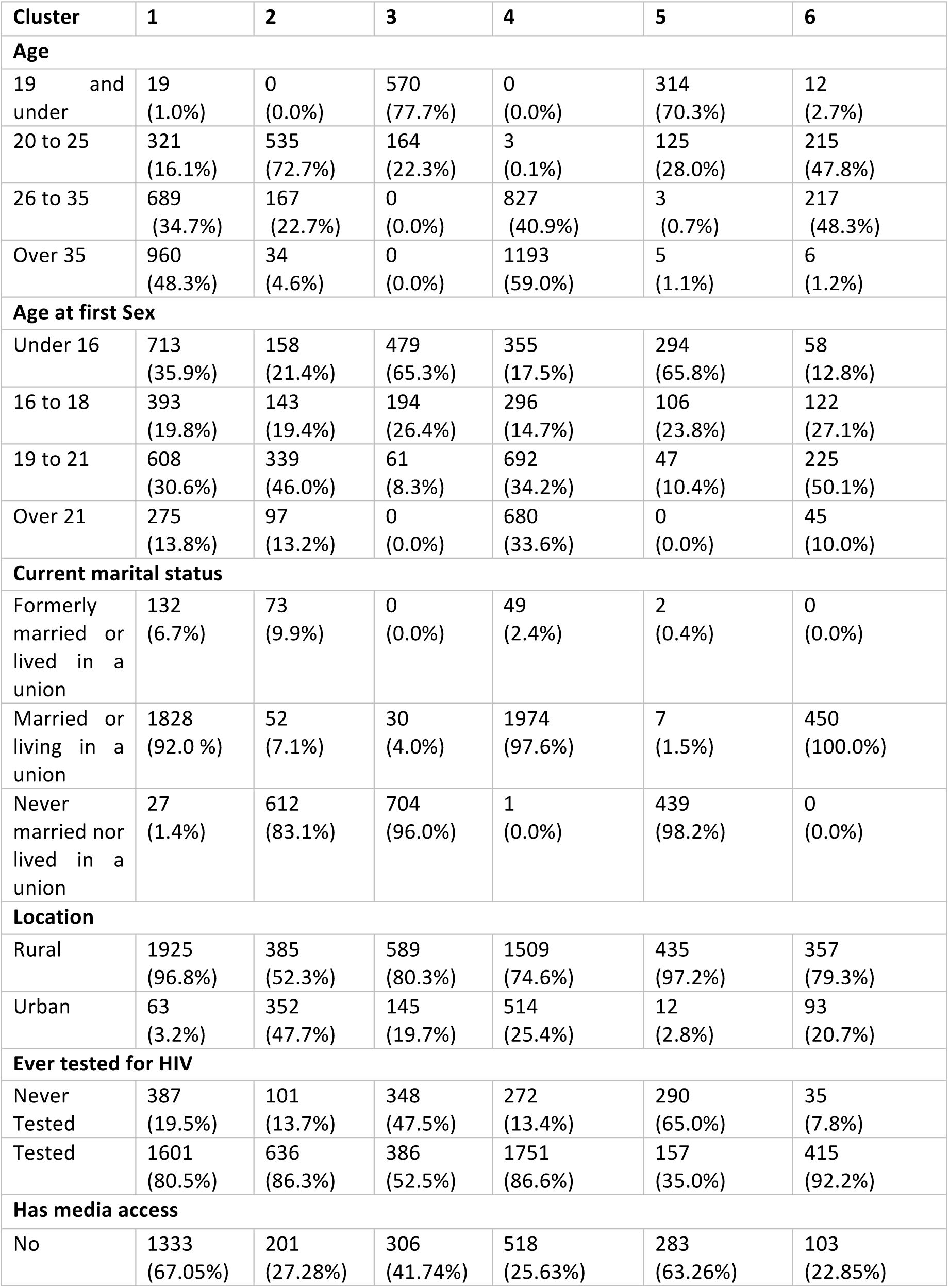
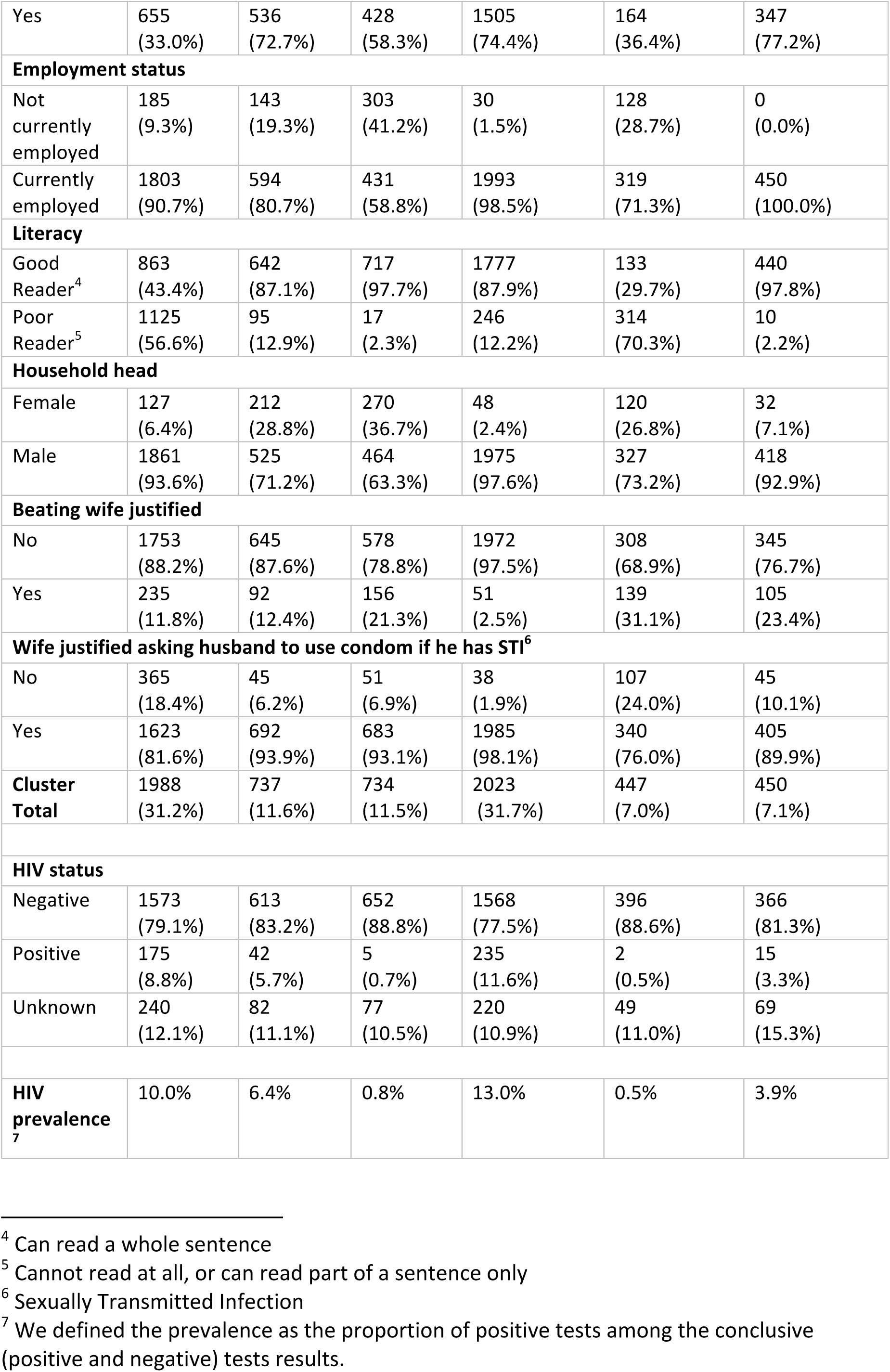
Clusters of men based on the 11 most important variables: age, age at first sex, marital status, location of residence, having ever been tested for HIV, media access, employment, literacy, sex of household head, wife beating justification and willingness to use condom. Data from 6,379 men were analyzed.

The clusters of men with the highest HIV prevalence were *Clusters 1* and *4* (10.0% in *Cluster 1;* 13.0% in *Cluster 4*). These clusters represented 31.2% and 31.7% of all men, respectively, and had a relatively high HIV testing uptake (80.5% in *Cluster 1*; 86.6% in *Cluster 4*). They both had a high proportion of older men (48.3% aged over 35 in *Cluster 1;* 59.0% in *Cluster 4*), who were mainly living in a union (92.0% in *Cluster 1;* 97.6% in *Cluster 4*). The household heads were also mostly men (93.6% in *Cluster 1;* 97.6% in *Cluster 4*). In contrast, *Cluster 4* was more urban than *Cluster 1* (25.4% vs 3.2%), and access to media in it was better (74.4% vs 33.0%). The cluster with the third highest HIV prevalence was *Cluster 2* (6.4%). This cluster had a high proportion of young men (72.7% between 20 and 25 years old), who were literate (87.1%) and had never lived in a union (83.1%). In comparison to *Cluster 2, Cluster 6*, which included mainly men who were literate (97.8%), aged between 20 and 35 (96.1%), had regular access to media (77.2%), and were living in a union (100.0%) and in rural settings (79.3%), had a lower HIV prevalence (3.9%) and a higher HIV testing uptake (92.2% in *Cluster 6* vs. 86.3% in *Cluster 2*).

**Figure S1** and **Table S5** present the percentage of each cluster of men in the Northern, Central and Southern regions of Malawi. The distribution of these clusters did not vary much between the three regions. It varied more between districts (**Table S6**). **Figure 3** presents the proportion of the six clusters in the districts of Chitipa (Northern), Salima (Central), Lilongwe (Central), Blantyre (Southern) and Chikwawa (Southern region). In both Central and Southern regions, the districts with higher HIV prevalence (Lilongwe; Blantyre) were characterized by a lower proportion of older men (over 35) living in rural areas, who were in a union and did not have regular access to media (22.8% of *Cluster 1* in Lilongwe vs 36.5% in Salima; 15.0% in Blantyre vs 30.7% in Chikwawa). Lilongwe and Blantyre also had a lower percentage of rural, illiterate adolescents with a low HIV prevalence (4.9% of *Cluster 5* in Lilongwe; 3.3% in Blantyre) than Salima and Chikwawa (14.5% of *Cluster 5* in Salima; 12.2% in Chikwawa). Higher HIV prevalence coincided also with a higher percentage of young, literate men, who had never lived in a union (18.5% of *Cluster 2* in Lilongwe vs 9.4% in Salima; 23.0% in Blantyre vs 7.8 % in Chikwawa), and a higher proportion of older (over 35) men, who were living in a union and had regular access to media (36.3% of *Cluster 4* in Lilongwe vs 21.0% in Salima; 42.0% in Blantyre vs 35.1% in Chikwawa). Both districts of the Southern region (Blantyre and Chikwawa) were characterized by a lower proportion of literate, married men, who were aged 20-35 and lived in rural settings (4.7% of *Cluster 6* in Blantyre; and 4.9% in Chikwawa vs 10.2%), compared to Chitipa (Northern region). In contrast, Chitipa was characterized by a lower percentage of rural, illiterate, adolescent men (5.1% of *Cluster 6*) than the rural districts of Southern and Central regions (12.2% in Chikwawa; 14.5% in Salima).

**Figure 3.**
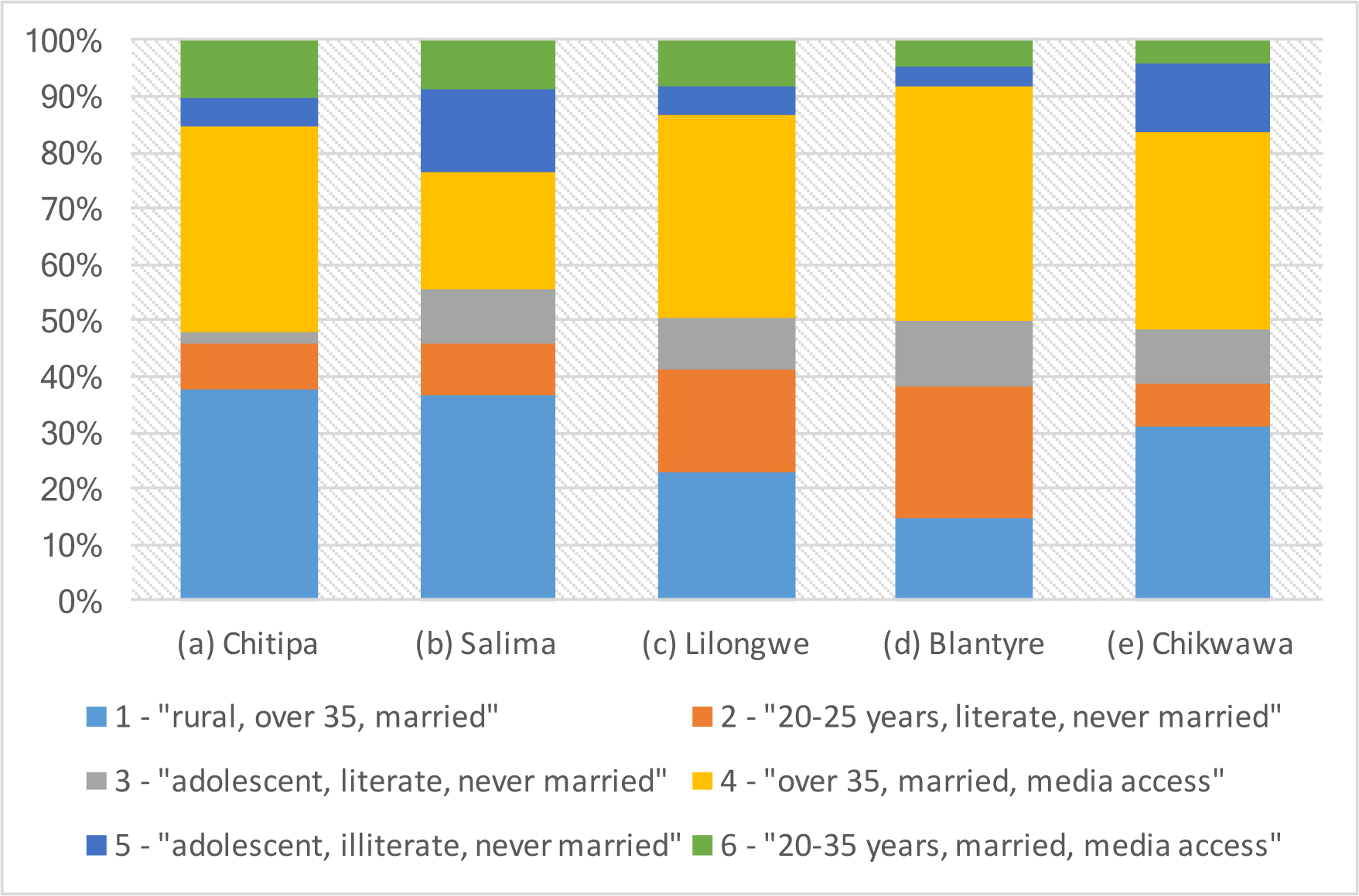
Proportion of the six male clusters in five selected districts of Malawi: Chitipa, (b) Salima, (c) Lilongwe, (d) Blantyre, (e) Chikwawa.

## Discussion

Using LCA, we categorized Malawian women into nine clusters and men into six clusters with common sociobehavioural characteristics. Among these clusters, we identified profiles of women and men who were most at risk of being HIV infected and less tested for HIV. Female household heads bore the heaviest burden of HIV infection, and rural adolescent girls had a significantly lower HIV testing uptake despite a relatively high HIV prevalence. Among men, older ones were the most affected by HIV, followed by young men who had never been married. Mapping the identified clusters across the regions and districts of Malawi showed that the clusters most vulnerable to HIV (higher prevalence and lower testing) were more frequent in the Southern region and the urban districts of the country.

The clusters of women with the highest HIV prevalence were characterized by an older age (over 25 years), living in a female-headed household and having lived formerly in a union. This finding is consistent with earlier studies that showed that women who were no longer in a union were most-at-risk of being HIV infected in Malawi[4]. The most common group of female household heads in Malawi are widows[16], and it is well possible that their husband died of AIDS. Indeed, the proportion of widows and orphans increased over time in the most HIV-affected countries[17]. Female household heads were also shown to live more frequently in poverty, requiring support from dedicated programmes[16,18].

The uptake of HIV testing in the identified clusters was generally consistent with the risk of being infected by HIV, with more persons being tested for HIV when HIV prevalence was high. Adolescents (<20 years) were tested much less frequently than adults[19–21]. The low HIV testing uptake among adolescents is concerning, because the proportion of young people (15-24) among all AIDS-related deaths in Malawi increased from 3.1% to 11.5% between 2004 and 2018[1]. In particular, rural adolescent girls, who have never lived in a union, were tested about half as much as older (20-35 years), rural, literate women, who were living in a union, and had a similar relatively high HIV prevalence. Yet, adolescent girls are a vulnerable cluster that often lack knowledge about HIV[19], tend to perceive themselves as being at low risk of HIV[22], and are prime targets of gender-based violence[23]. These observations confirm the need to accelerate tailored HIV prevention programming among adolescents in general, and specifically among rural adolescent girls[24].

Men tended to have a lower HIV prevalence than their female counterparts, confirming the gender disparity in HIV infection in SSA[25,26]. Men were also less often tested for HIV, which could be influenced by harmful masculinity stereotypes and cultural norms that inhibit men from seeking care[27,28]. It may also result from the focus of HIV prevention in recent years on women and girls, and HIV testing being routinely performed in antenatal care. UNAIDS’ report released in 2017[29] highlighted the low utilisation of health services by men and qualified it as a *blind spot* in the response to HIV. Men have therefore a higher risk of HIV transmission to their partners, and eventually a higher mortality rate due to AIDS-related illnesses, affecting directly their families.

HIV prevalence was associated with the age and partnership status of men across the clusters. Older men (>35 years) had the highest HIV prevalence regardless of their place of residence, literacy, or access to media. Compared to 2016, HIV incidence in Malawi was nearly three times higher in year 2000[1], which can explain the higher risk of HIV acquisition among men who were young adults at that time. The current lower willingness to condom utilisation among rural, less educated, older and married men[30,31] may also contribute to higher risk of recent HIV acquisition. Among young men, living in a union and in a rural setting was associated with a reduced risk of HIV. This finding is in agreement with a recent study performed in South Africa, where single never married men had a high risk of being HIV infected, which may be associated with a higher frequency of less stable relations and multiple sexual partners[32].

Clusters of women and men with high prevalence of HIV and low HIV testing coverage were more frequent in urban areas of Malawi. Palk and Blower[7] showed that urban areas of Malawi also had a higher density of individuals with a high number of lifetime sexual partners. Women and men in the mostly rural Northern region were generally well tested. Our finding is in agreement with a recent study, which also reported a regional variance of HIV testing uptake in Malawi, with a better testing uptake in the North, although the latter study focused on men only[13].

Our work has some limitations. The analysis was based on a limited number of factors that were selected by the researchers based on prior knowledge from the available literature. In order to avoid the subjective selection of the variables in the future, we are working in parallel on an automatic method to select the most important variables to predict individual HIV status. Our preliminary results showed that most of the variables that we selected in this study were highly relevant to predict HIV. Our analysis allowed us to identify clusters of men and women with associated probabilities of being HIV infected and tested for HIV, but the cross-sectional nature of our analysis does not allow us to draw causal relations. There are other methods, such as Bayesian network analyses, that attempt to determine the possible causal pathways between various risk factors and HIV[33]. The data we used are also potentially biased because they are self-reported (ex. more educated people may be less keen to admit gender-based violence or non-willingness to use condoms).

In conclusion, using LCA to identify and map vulnerable clusters of women and men can provide key information to policy makers, enabling them to strategically design tailored support and prevention programs that target people who are most in need. Our results clearly demonstrated that rural adolescent girls, young single men, and married men aged over 35 years have a particularly high risk of being infected by HIV. Efforts should be made to increase the coverage of HIV testing in particular among these sub-populations.

## Data Availability

The data generated by our analysis and the R code we developed for this work are available online.

https://gitlab.com/AzizaM/dhs_malawi_lca

## Acknowledgement

We acknowledge the support of the Swiss National Science Foundation (SNF professorship grant n° 163878 to O Keiser) who funded this study. We thank Isotta Triulzi for helpful discussions and comments.

